# Sensitivity of the African Neuropsychology Battery Memory Subtests and Learning Slopes in Discriminating APOE 4 and Amyloid Pathology in Adult Individuals in the Democratic Republic of Congo

**DOI:** 10.1101/2023.10.05.23296598

**Authors:** Jean Ikanga, Sarah D. Patrick, Megan Schwinne, Saranya Sundaram Patel, Emmanuel Epenge, Guy Gikelekele, Nathan Tshengele, Immaculee Kavugho, Samuel Mampunza, Kevin E. Yarasheski, Charlotte E. Teunissen, Anthony Stringer, Allan Levey, Julio C. Rojas, Brandon Chan, Argentina Lario Lago, Joel H. Kramer, Adam L. Boxer, Andreas Jeromin, Alvaro Alonso, Robert J. Spencer

## Abstract

**Background:** The current study examined the sensitivity of two memory subtests and their corresponding learning slope metrics derived from the African Neuropsychology Battery (ANB) to detect amyloid pathology and APOEε4 status in adults from Kinshasa, the Democratic Republic of the Congo.

**Methods:** 85 participants were classified for the presence of β-amyloid pathology and based on allelic presence of APOEε4. All participants were screened using CSID and AQ, underwent verbal and visuospatial memory testing from ANB, and provided blood samples for plasma Aβ_42_, Aβ_40_, and APOE proteotype. Pearson correlation, linear and logistic regression were conducted to compare amyloid pathology and APOEε4 status with derived learning scores, including initial learning, raw learning score, learning over trials, and learning ratio.

**Results:** Our sample included 35 amyloid positive and 44 amyloid negative individuals as well as 42 without and 39 with APOEε4. All ROC AUC ranges for the prediction of amyloid pathology based on learning scores were low, ranging between 0.56-0.70 (95% CI ranging from 0.44-0.82). The sensitivity of all the scores ranged between 54.3-88.6, with some learning metrics demonstrating good sensitivity. Regarding APOEε4 prediction, all AUC values ranged between 0.60-0.69, with all sensitivity measures ranging between 53.8-89.7. There were minimal differences in the AUC values across learning slope metrics, largely due to the lack of ceiling effects in this sample.

**Discussion:** This study demonstrates that some ANB memory subtests and learning slope metrics can discriminate those that are normal from those with amyloid pathology and those with and without APOEε4, consistent with findings reported in Western populations.

## INTRODUCTION

Problems learning and remembering are common across neurodegenerative conditions and can contribute to declines in everyday functioning^1^. In clinical evaluations, learning and memory are frequently evaluated using learning tasks, in which examinees are presented with verbal or visual stimuli over several trials and asked to learn, and then subsequently recall, the information after a delay. The primary test scores include the total learning score (i.e., the total amount of information learned over the repeated trials) and the short- and long-delay memory scores (i.e., the amount of information an examinee recalled after delays). These scores are associated with amyloid and tau biomarkers^2,3^ and brain volume,^4^ aiding in clinically relevant outcomes such as the early detection of Alzheimer’s disease (AD).^5,6^

Clinicians often use supplementary scores to parse apart the cognitive process test takers use when completing these memory tasks. One of these scores is learning slope, which refers to the rate at which the examinee improves throughout the learning trials. Healthy adults are expected to benefit from repeated exposure to information; however, patients with neurological conditions, such as AD, Korsakoff’s syndrome, or amnestic mild cognitive impairment, commonly demonstrate a lack of improvement from practice or repetition,^7–10^ or more formally known as a “shallow” or “flat” learning slope.

Learning slope calculations can be applied to practically any measure involving repeated administration of to-be-learned stimuli and has been used with a variety of visual and verbal list learning tasks, including the California Verbal Learning Test (CVLT),^11^ Hopkins Verbal Learning Test-Revised (HVLT-R),^12^ the Rey Auditory Verbal Learning Test (RAVLT),^13^ Repeatable Battery for the Assessment of Neuropsychological Status (RBANS),^14,15^ and Brief Visual Memory Test-Revised (BVMT-R).^16^ The most used learning slope metric, the raw learning slope (RLS), is usually calculated by subtracting the first learning trial score from the final learning trial score. Unfortunately, the interpretability of RLS is constrained by ceiling effects for examinees who perform well on the first trial because there is limited opportunity to learn new information on subsequent trials. For example, an examinee who learns 2 words on a 16-word list can achieve an RLS score of 14, whereas an examinee who learned 10 words on the first trial can obtain a maximum RLS of only 6. Spencer and colleagues addressed the drawbacks to RLS with their proposal of a new method, the learning ratio (LR), which directly addresses the initial learning score.^17^ This score mathematically accounts for initial learning trial performance by calculating information learned as a percentage of the amount of information left to be learned.

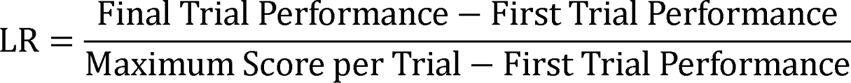

Although the RLS score or some close derivation thereof is usually depicted in test manuals, the LR score has a growing number of normative datasets for several learning and memory tests, including the RAVLT,^18^ HVLT-R and BVMT-R,^19^ RBANS,^20^ and African Neuropsychology Battery (ANB),^21^ providing clinicians with anchors for evaluating performance. By eliminating confounds from ceiling effects, the LR may result in stronger associations and predictive ability for clinically relevant outcomes than the RLS, which typically produces weak to null findings when evaluated in the context of demographic characteristics and other performance characteristics.^10^ When both scores are directly compared, LR performs equal to or better than RLS for correlations with memory tests,^3,17,18,22,23^ diagnostic discrimination,^3,18,23^ as well as other known risk factors for AD, including hippocampal volume, AD-specific biomarkers, and APOEε4 status.^22–24^ Thus, the LR score is a relatively new but promising metric that warrants additional examination, particularly in the context of detecting AD using non-invasive assessment methods.

AD is associated with brain-based changes including the presence of abnormally high amyloid beta plaque deposition and pathologic tau aggregations.^25^ These physiological changes can be detected years before the development of clinical AD symptoms.^26^ Tau burden^27^ and amyloid deposition^28^ consistently correlate with performance on memory measures. Amyloid beta and tau burden are typically measured using positron emission tomography (PET) scans, cerebrospinal fluid (CSF), or blood plasma. Unfortunately, both PET and CSF are costly, invasive, and inaccessible to many patients. Such practical concerns are most important in communities with limited access to medical and financial resources, such as in the Democratic Republic of Congo (DRC). In contrast, blood plasma is a faster, cheaper, and more accessible alternative.^29,30^ Plasma P-tau181 values are correlated with increased tau PET measurements and have shown promise in discriminating between AD and non-AD degenerative disease, reflecting severity of AD across the AD continuum, and progression to AD.^29^

Administering paper-and-pencil cognitive tests is cheaper, more portable/accessible, and less invasive than amyloid or tau PET imaging, cerebrospinal fluid (CSF) sampling and analysis, and more recently blood sampling for plasma biomarker analysis. Research on memory test process scores and AD biomarkers indicate that worse LR scores are associated with greater cerebral beta-amyloid deposition (PET) and APOEε4 positivity,^22^ though these scores have not yet been evaluated in the context of blood plasma biomarker status for the ANB. Given the practical advantages of the LR score, any indications of subtle cognitive inefficiencies that are associated with AD-specific plasma biomarkers and downstream cognitive changes suggest that this score may be a worthwhile metric for identifying early indications of AD pathology.

This study examined two memory subtests and their corresponding learning slope metrics from the ANB to aid in detecting amyloid pathology and APOEε4 allele presence.^25^ We hypothesized that ANB memory and learning slope scores would have adequate to good test receiver operator characteristics (ROC; e.g., area under the curve, specificity, sensitivity) to classify patients into plasma amyloid positive or negative. Based on previous studies,^22^ we predicted that the LR score would bear a stronger relationship to amyloid pathology and APOEε4 allele than would the other learning slopes.

## METHODS

### Study population

Participants were selected from our previous study,^29^ a matched case-control study to identify risk factors of AD in Sub-Saharan Africa (SSA). Participants were at least 65 years or older, had a family member or close friend to serve as an informant, and fluent in French or Lingala. Participants were excluded if they had history of schizophrenia, neurological, or other or medical conditions potentially affecting the CNS. In the absence of established diagnostic criteria for AD in SSA, we used two screening measures with high sensitivity and specificity for identifying individuals with dementia in Western cohorts, the Alzheimer’s Questionnaire (AQ)^31^ and the Community Screening Instrument for Dementia (CSID),^32^ which evaluate clinical symptoms of dementia. The AQ distinguishes between those with AD from healthy controls.^31^ The CSID Questionnaire has been extensively used in many international and SSA dementia studies.^32^

Based on cognitive and functional deficits for Diagnostic and Statistical Manual of Mental Disorders, Fifth Edition, Text Revision (DSM-5-TR) diagnostic criteria,^33^ we used Brazzaville cut-offs of CSID, the closest city from Kinshasa, to classify participants.^34^ Similar to our prior study,^35^ participants were classified using CSID and AQ scores (see Figure 1), which yielded 4 groups: major neurocognitive disorder/dementia, mild neurocognitive disorder (MND), subjective cognitive impairment, and healthy control (HC), i.e., normal cognition.

**Figure 1.**
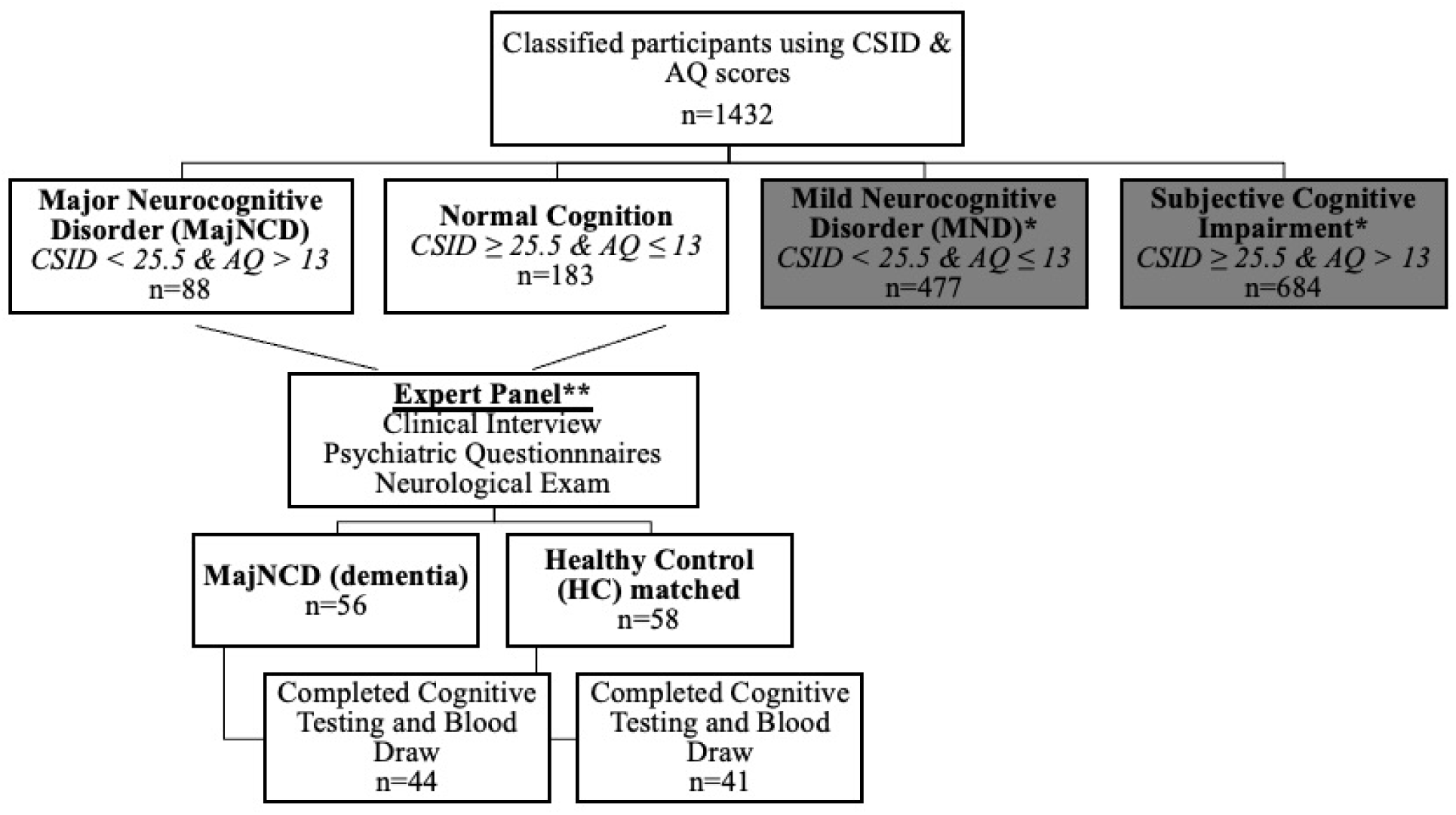
Flow diagram of participant classifications using the CSI-D and the AQ in the current study. **Abbreviations**: CSID (Community Screening Instruments for Dementia); AQ (Alzheimer's Questionnaire); MajNCD (major neurocognitive disorder); HC (heathy controls); MND (mild neurocognitive disorder).

A panel consisting of a neurologist, psychiatrist and neuropsychologist reviewed screening tests, clinical interview, and neurological examination of subjects. 56 individuals were confirmed with a diagnosis of dementia and 58 were considered HC. Participants were matched based on age, education, and gender. Plasma biomarkers were obtained for 85 subjects (75%), resulting in a final sample of 44 dementia and 41 HC. The remaining 29 subjects refused to provide blood samples. Written informed consent was obtained prior to participants’ undergoing any study procedures. Participants were financially compensated for their time. The informed consent document and all research procedures were approved by the Ethics Committee/Institutional Review Boards of the University of Kinshasa.

### Procedure

Participants underwent a comprehensive clinical evaluation, including cognitive testing, self-report questionnaires, and standard psychiatric and neurological evaluations to be diagnosed with dementia or to be considered as HC by an expert panel [neurologist (EE), psychiatrist (GG), and neuropsychologist (JI)]. Subjects were interviewed to obtain demographic, socioeconomic, and medical history and subsequently administered cognitive testing with ANB subtests. Afterwards, blood samples were obtained at Medical Center of Kinshasa (CMK) by a phlebotomist. Sample collection protocol and quantification of fluid biomarkers are presented below.

### Measures

#### Plasma biomarkers

Blood samples were drawn in the CMK blood laboratory by venipuncture into dipotassium ethylene diamine tetra acetic acid (K_2_ EDTA) tubes. Samples were centrifuged within 15 minutes, and 5 mL of plasma was aliquoted into 0.5 mL polypropylene tubes and stored initially at −20° C for less than a week and then moved to a −80 °C freezer for longer term storage at a CMK laboratory.^36^ These aliquots were shipped frozen on dry ice to Emory University and APOE isoform-specific peptides were analyzed at C_2_N Diagnostics (St. Louis, MO) as described.^37^ We used LC-MS/MS to detect and identify the APOE isoform-specific peptides (ε2, ε3, ε4), with the purpose being to classify participants into APOEε carriers and non-carriers Plasma biomarker concentrations were measured using commercially available Neurology 4-PLEX E (Aβ40, Aβ42), P-tau181 (P-tau181 v2; l) Quanterix kits for the Simoa HD-X platform (Billerica, MA) at the University of San Francisco. P-tau217 was measured using the proprietary ALZpath P-tau-217 CARe Advantage kit (lot #MAB231122, ALZpath, Inc.) for the Simoa HD-X platform. The instrument operator was blinded to clinical variables. All analytes were measured in duplicate. For Aβ_40_ and Aβ_42_, all samples were measured above the lower limit of quantification (LLOQ) of 1.02 pg/mL and 0.378 pg/mL. The average coefficient of variation (CV) for Aβ_40_ and Aβ_42_ were 6.0% and 6.5%. For P-tau181, all samples were measured above the kit lower limit of quantification (LLOQ) of 0.085 pg/mL, with an average CV of 11.6%. For P-tau217 the LLOQ was 0.024 pg/mL and the average CV was 19.8%.

In the absence of a gold standard and universal cut-offs available across different assays measuring plasma β-amyloid or tau, we have chosen diagnostics β-amyloid (A+ = Aβ_42/40_ < 0.056) cutoff.^38^ Based on this cutoff, we classified participants into either normal (A-) and amyloid pathology (A+) groups.

#### Cognition

Cognitive functioning was evaluated using the learning and memory subtests from the ANB, including the: African List Memory Test (ALMT; verbal learning and memory; long delay free recall correct score) and African Visuospatial Memory Test (AVMT; visuospatial memory; long delay recall correct score). The ANB has been shown to have good psychometric properties in evaluating effects of aging and neurological disease alongside providing culturally and linguistically appropriate neuropsychological measures for SSA countries.^35^

##### African List Memory Test (ALMT)

The ALMT is a measure of unstructured verbal learning and memory. It consists of 12 words from 4 semantic categories (body parts, means of transportation, animals, and food). The list is read at the rate of one word per 3 seconds across 3 learning trials. Examinees are asked to recall words from the target list immediately following the presentation of the list (raw scores ranging from 0 to 12). A second 12-word interference list is then presented after the third learning trial, followed by the short-delayed recall, and then a long-delay free recall trial approximately 20 minutes later.^39^

##### African Visuospatial Memory Test (AVMT)

The AVMT is measure of visuospatial learning and memory. This task involves encoding and retaining traditional cultural symbols found in the arts, including woodcarving, textiles, and prints, inherent to many SSA countries.

The examinee is first presented with a page of 4 symbols organized in a 2×2 matrix. The page is displayed for 10 seconds, after which examinees were asked to reproduce (on a blank sheet of paper) as many of the symbols, in their correct location on the page. For each of the 4 individual symbols, scores range from 0 to 5 yielding a total score (accuracy of the drawing and location) from 0 to 20 for each trial.^39^

### Calculation of Initial Learning and Learning Slopes

#### Initial learning

Initial learning was assessed using Trial 1 score for each measure. A weighted initial learning score was calculated by adjusting the point values for each test by the number of items, with the test awarding the most points per trial - AVMT (20 points) – serving as the standard.

Because the highest score of items in ANB memory tests is 20, we made 20 the total for each trial to match the maximum total of 20 in AVMT. The weighted average is adjusted for the number of items for each test (e.g., 1.67 * 12 = 20). Therefore, ALMT contains 12 possible points that are multiplied by 1.67 so the total approximated the 20 points of AVMT.

We used the following formula:

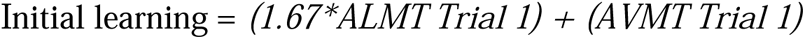

#### Learning Slope

For learning slope we calculated raw learning score (RLS), LOT (learning over trials) (modified from Morrison et al., 2018), learning ratio (LR) (Spencer et al., 2020), and total LR. See below for equations.

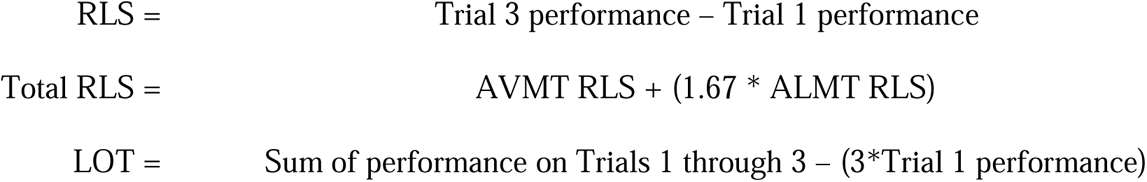

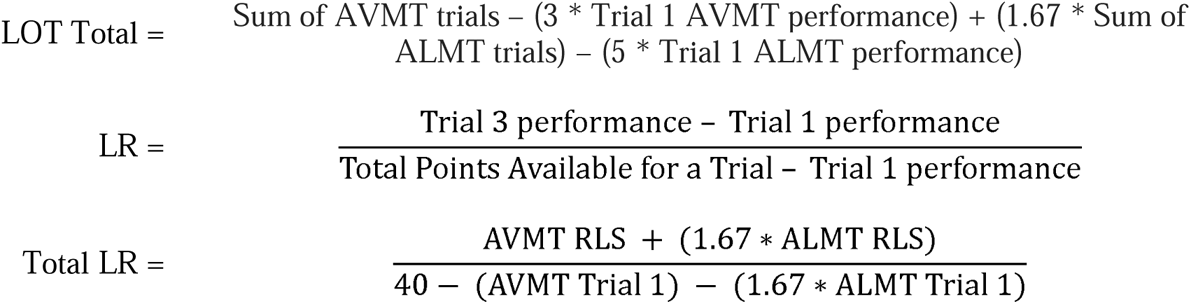

### Statistical Analyses

Statistical analyses were performed using SAS statistical software.^33^ Pearson correlations were calculated between individual learning slope performances and ANB memory measures. Linear regression was used to assess between-group differences of various learning slope scores (RLS, LOT, and LR) in which amyloid pathology and APOEε4 status served as predictor variables. These analyses controlled for demographic characteristics (i.e., age, education, gender). We used receiver operating characteristic (ROC) curve analyses to calculate areas under curve (AUCs) to describe diagnostic accuracy of amyloid pathology based on screening tests (CSID, AQ), ANB memory scores, and learning slope metrics (RLS, LOT, LR). We divided the participants based on amyloid positivity or negativity based on our previous cutoffs for amyloid pathology. We used Hosmer and colleagues ROC-AUC categories,^40^ which considered the value of <0.600 as “failure”, values between 0.600 and 0.699 as “poor”, values between 0.700 and 0.799 as “fair”, values between 0.800 and 0.899 as “good”, and values 0.900 or greater as “excellent”. Cutoff scores for screening tests and ANB learning slopes were determined based on optimal sensitivity and specificity for detecting the presence of β-amyloid pathology. We obtained Youden’s J indices (sensitivity + specificity – 100) for each plasma biomarker. We estimated the predicted value of the measure and then the cutoffs. We also calculated the Cohen’s d as follows:

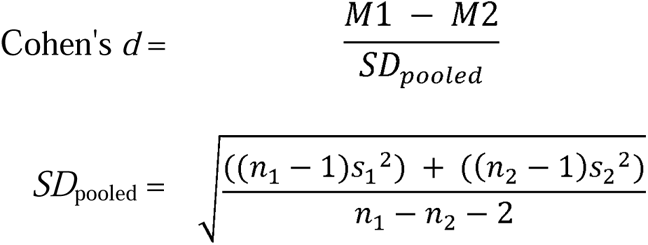

## RESULTS

### Demographic, Cognitive, and Clinical Characteristics of Sample

Nearly half of participants had amyloid pathology (44%), and nearly half were APOEε4 positive (49%). Demographic data, neuropsychological scores, and plasma biomarker characteristics stratified by amyloid pathology are presented in Table 1. Groups did not significantly differ between those with and without amyloid pathology with regard to demographics (e.g., age, education, sex). The groups did not differ with respect to cognition after controlling for age, education, and gender. Based on Cohen’s d results, AQ total score, delayed recall total, RLS total, and LOT total had at least moderate effect sizes, while the LR total score had a large effect size. Regarding subtest scores, most of the learning slope scores from each of the subtests ranged from small to moderate effect sizes.

**Table 1.**
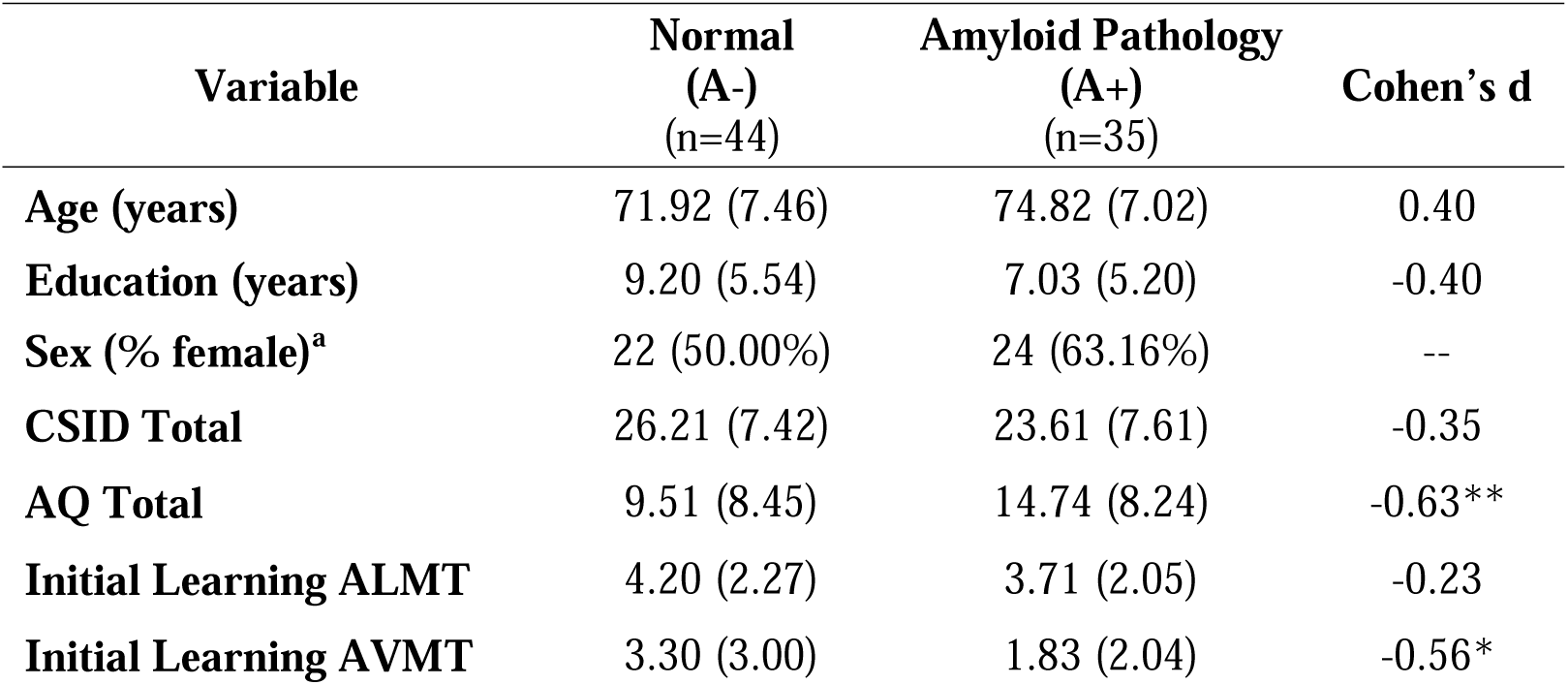
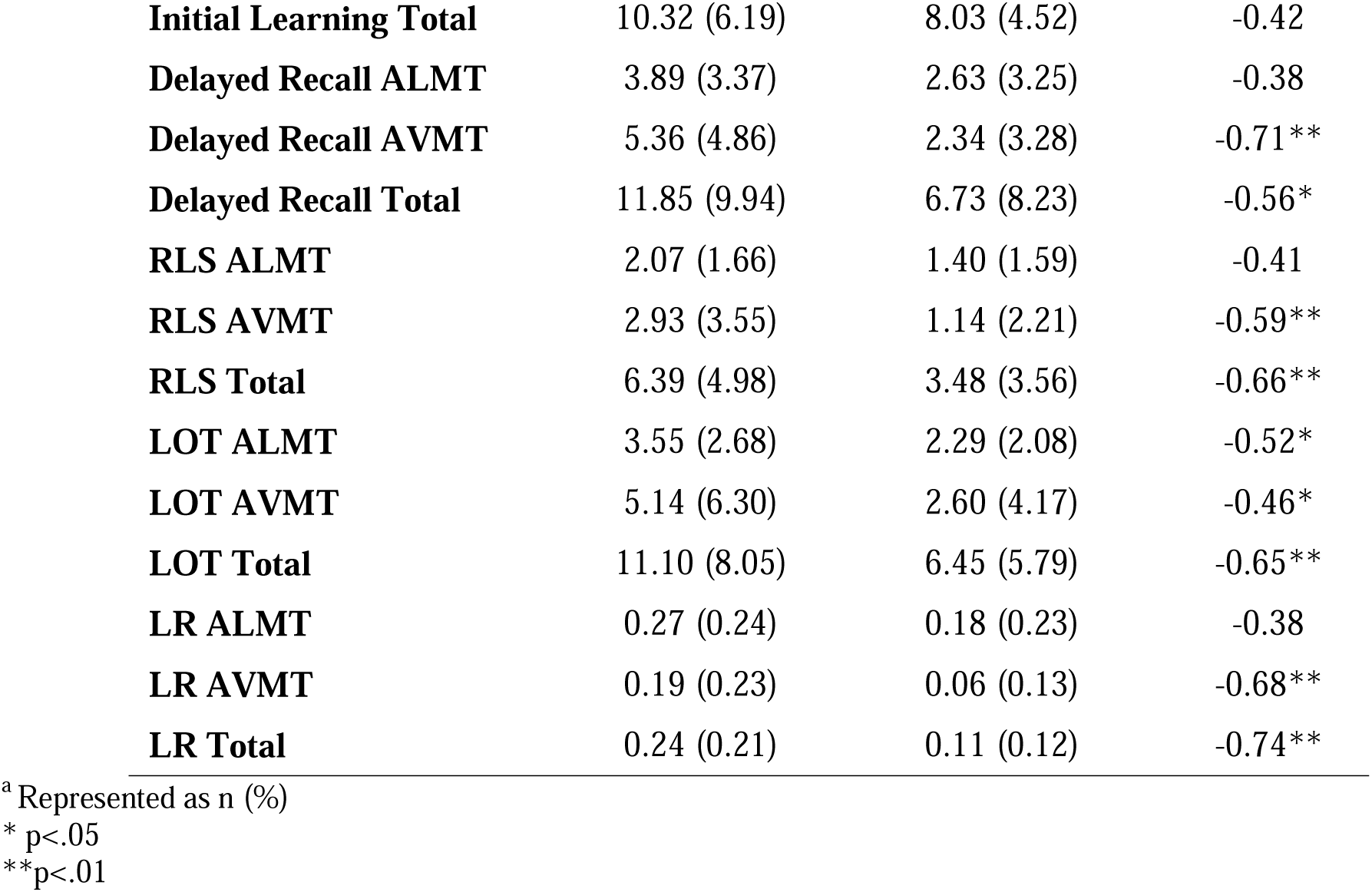
Demographic, neuropsychological, and behavioral variables stratified by amyloid pathology with the Cohen’s d.

Regarding APOEε4 status, some significant group differences were observed. Specifically, those without APOEε4 performed significantly better than those with the ε4 allele across cognitive screening measures and tasks of verbal and visual learning and memory. For the primary test scores, initial learning for visuospatial information had a small effect size, while delayed recall, RLS, and LOT scores for visuospatial information had moderate effect sizes. The learning slope metrics were comparable with initial learning total score having a small effect size alongside mostly moderate effect sizes for RLS, LOT, and delayed recall scores (see Table 2).

**Table 2.** Demographic, neuropsychological, and behavioral variables stratified by APOEε4status with Cohen’s D.

| **Variable** | **APOε4-**<br>**(n=42)** | **APOε4+**<br>**(n=39)** | **Cohen's d** |
| --- | --- | --- | --- |
| **Age (years)** | 71.85 (7.06) | 74.10 (8.20) | 0.29 |
| **Education (years)** | 8.21 (5.44) | 8.54 (5.29) | 0.06 |
| **Sex (% female)<sup>a</sup>** | 27 (62.79%) | 20 (48.78%) |  |
| **CSID Total** | 27.01 (7.86) | 23.03 (6.91) | -0.54* |
| **AQ Total** | 9.54 (8.45) | 14.56 (8.48) | 0.59** |
| **Initial Learning ALMT** | 4.50 (2.06) | 3.49 (2.22) | -0.47* |
| **Initial Learning AVMT** | 3.07 (2.77) | 2.23 (2.57) | -0.31 |
| **Initial Learning Total** | 10.59 (5.49) | 8.05 (5.48) | -0.46* |
| **Delayed Recall ALMT** | 4.45 (3.39) | 2.18 (3.02) | -0.71** |
| **Delayed Recall AVMT** | 5.43 (4.52) | 2.95 (4.53) | -0.55* |
| **Delayed Recall Total** | 12.86 (9.71) | 6.59 (9.00) | -0.67** |
| **RLS ALMT** | 2.10 (1.86) | 1.44 (1.48) | -0.39 |
| **RLS AVMT** | 3.10 (3.41) | 1.44 (3.32) | -0.49* |
| **RLS Total** | 6.59 (5.00) | 3.83 (4.83) | -0.56* |
| **LOT ALMT** | 3.38 (2.70) | 2.46 (2.39) | -0.36 |
| **LOT AVMT** | 5.64 (5.88) | 2.54 (5.23) | -0.56* |
| **LOT Total** | 11.33 (7.83) | 6.68 (7.10) | -0.58** |
| **LR ALMT** | 0.29 (0.27) | 0.17 (0.21) | -0.33* |
| **LR AVMT** | 0.19 (0.21) | 0.09 (0.20) | -0.05* |
| **LR Total** | 0.24 (0.19) | 0.13 (0.19) | -0.26* |

<sup>a</sup> Represented as n (%)
\* p<.05
\*\*p<.01

We calculated the sensitivity and specificity of screening and memory subtests to predict amyloid positivity or negativity and the presence of APOEε4 allele (see Table 3). In predicting amyloid pathology, sensitivity of cognitive tests varies between 56.8 (CSID) to 82.9 (Delayed AVMT). The sensitivity of cognitive tests to predict APOEε4 allele was between 60.5 (Aβ_42/40_) and 74.4 (Delayed AVMT).

**Table 3.** Sensitivity and Specificity of Cognitive Tests to Predict A+ or APOEε4+ status.

| **Test**<br>(Cognitive Impairment Threshold) |  | **Aβ<sub>42/40</sub>** |  |  | **APOE ε4** |  |  |
| --- | --- | --- | --- | --- | --- | --- | --- |
|  |  | **High**<br>**Aβ<sub>42/40</sub>**<br>**(A-)** | **Low**<br>**Aβ<sub>42/40</sub>**<br>**(A+)** | **SE / SP**<br>**(%)** | **APOε4 –** | **APOε4 +** | **SE / SP**<br>**(%)** |
| **CSID Screening** | ≥25.5 (HC) | 25 | 16 | 59.5. /56.8 | 26 | 15 | 65.0 / 62.5 |
|  | <25.5 (Dem) | 17 | 21 |  | 14 | 25 |  |
| **AQ Screening** | <13 (HC) | 24 | 13 | 55.8 / 65.8 | 25 | 12 | 61.0 / 70.7 |
| | $\geq 13$ (Dem) | 19 | 25 | | 16 | 29 | |
| **Delayed Recall ALMT** | $\geq 5$ (HC) | 23 | 13 | 52.3 / 62.9 | 26 | 11 | 61.9 / 71.8 |
| | $< 5$ (Dem) | 21 | 22 | | 16 | 28 | |
| **Delayed Recall AVMT** | $\geq 5$ (HC) | 25 | 6 | 56.8 / 82.9 | 23 | 10 | 54.8 / 74.4 |
| | $< 5$ (Dem) | 19 | 29 | | 19 | 29 | |
| **A$\beta_{42/40}$** | $> 0.056$ (HC) | | | | 26 | 17 | 60.5 / 60.5 |
| | $\leq 0.056$ (Dem) | | | | 15 | 23 | |

\*CIP = cognitive impairment

We calculated Pearson correlation coefficients to examine the relationship between total learning slope metrics, initial learning, and delayed recall (See Table 4). Among the learning slope options, LR had the strongest correlations with both initial and delayed recall. The learning slope metrics generally had high correlations with each other.

**Table 4:** Pearson correlations between total learning slope performances.

| Variable | Initial Learning Total | RLS Total | LOT Total | LR Total |
| --- | --- | --- | --- | --- |
| **RLS Total** | 0.18 (0.056) | - |  |  |
| **LOT Total** | 0.20 (0.036) | 0.94 (<.0001) | - |  |
| **LR Total** | 0.39 (<.0001) | 0.95 (<.0001) | 0.89 (<.0001) | - |
| **Delayed Recall Total** | 0.82 (<.0001) | 0.48 (<.0001) | 0.51 (<.0001) | 0.63 (<.0001) |

### Analyses of ROC-AUC, Cutoffs, and Sensitivity/Specificity

Tables 5 and 6 display the ROC-AUC values for the screening measures, ANB memory subtests, learning slopes, and total scores when differentiating individuals between groups with amyloid pathology and APOEε4 status. Our results showed that all AUC/ROC ranges fell between 0.56-0.70 (95% *CIs* ranging from 0.44-0.82) and the sensitivity varied between 51.4-88.6.

**Table 5.** Receiver operating characteristic area under the curve, cut scores, and sensitivity/specificity with differentiating amyloid biomarker groups for screening tests, ANB memory subtests, and learning slopes across the total sample.

| **Variable** | **AUC**<br>**(95% CI)** | **Cut Score** | **Sensitivity/**<br>**Specificity** |
| --- | --- | --- | --- |
| **CSID Total** | 0.61<br>(0.48,0.74) | 27.33 | 64.9 / 54.8 |
| **AQ Total** | 0.68<br>(0.57,0.80) | 6.00 | 76.3 / 53.5 |
| **Delayed Recall ALMT** | 0.61<br>(0.49,0.73) | 5.00 | 74.3 / 45.5 |
| **Delayed Recall AVMT** | 0.68<br>(0.56,0.79) | 4.00 | 77.1 / 63.6 |
| **Delayed Recall Total** | 0.65<br>(0.53,0.77) | 12.02 | 68.6 / 54.5 |
| **Initial Learning ALMT** | 0.56<br>(0.44,0.69) | 6.00 | 82.9 / 27.3 |
| **Initial Learning AVMT** | 0.65<br>(0.53,0.77) | 3.00 | 77.1 / 50.0 |
| **Total Initial Learning** | 0.63<br>(0.50,0.75) | 10.35 | 77.1 / 43.2 |
| **RLS ALMT** | 0.62<br>(0.50,0.75) | 3.00 | 80.0 / 43.2 |
| **RLS AVMT** | 0.67<br>(0.55,0.78) | 3.00 | 77.1 / 50.0 |
| **RLS Total** | 0.70<br>(0.58,0.82) | 2.67 | 54.3 / 81.8 |
| **LOT ALMT** | 0.64<br>(0.52,0.76) | 4.00 | 88.6 / 34.1 |
| **LOT AVMT** | 0.62<br>(0.49,0.74) | 6.00 | 77.1 / 36.4 |
| **LOT Total** | 0.69<br>(0.57,0.81) | 5.37 | 51.4 / 79.5 |
| **LR ALMT** | 0.62<br>(0.50,0.75) | 0.33 | 80.0 / 38.6 |
| **LR AVMT** | 0.68<br>(0.56,0.80) | 0.17 | 85.7 / 45.5 |
| **LR Total** | 0.70<br>(0.58,0.82) | 0.10 | 60.0 / 72.7 |

**Table 6.** Receiver operating characteristic area under the curve, cut scores, and sensitivity/specificity with differentiating APOEε4+ for cognitive screening tests, ANB memory subtests, and learning slopes across the total sample.

| **Variable** | **AUC**<br>**(95% CI)** | **Cut Score** | **Sensitivity/**<br>**Specificity** |
| --- | --- | --- | --- |
| **CSID Total** | 0.67<br>(0.55, 0.80) | 27.33 | 70.0 / 60.0 |
| **AQ Total** | 0.65<br>(0.53, 0.77) | 9.00 | 70.7 / 61.0 |
| **Delayed Recall ALMT** | 0.68<br>(0.57, 0.80) | 5.00 | 71.8 / 61.9 |
| **Delayed Recall AVMT** | 0.68<br>(0.57, 0.80) | 5.00 | 82.1 / 50.0 |
| **Delayed Recall Total** | 0.70<br>(0.58, 0.81) | 10.01 | 71.8 / 64.3 |
| **Initial Learning ALMT** | 0.64<br>(0.52, 0.76) | 4.00 | 53.8 / 81.0 |
| **Initial Learning AVMT** | 0.60<br>(0.48, 0.72) | 5.00 | 89.7 / 31.0 |
| **Initial Learning Total** | 0.66<br>(0.53, 0.78) | 6.68 | 43.6 / 81.0 |
| **RLS ALMT** | 0.63<br>(0.51, 0.75) | 3.00 | 82.1 / 47.6 |
| **RLS AVMT** | 0.69<br>(0.57, 0.80) | 2.00 | 69.2 / 61.9 |
| **RLS Total** | 0.68<br>(0.56, 0.79) | 5.68 | 74.4 / 50.0 |
| **LOT ALMT** | 0.61<br>(0.48, 0.73) | 5.00 | 84.6 / 33.3 |
| **LOT AVMT** | 0.69<br>(0.58, 0.81) | 1.00 | 53.8 / 73.8 |
| **LOT Total** | 0.69<br>(0.57, 0.81) | 4.37 | 46.2 / 83.3 |
| **LR ALMT** | 0.66<br>(0.54, 0.78) | 0.33 | 87.2 / 45.2 |
| **LR AVMT** | 0.69<br>(0.57, 0.81) | 0.06 | 69.2 / 61.9 |
| **LR Total** | 0.69<br>(0.57, 0.80) | 0.11 | 56.4 / 71.4 |

Regarding the APOEε4, all ROC/AUC values ranged between 0.60-0.70. With regards to total scores, the sensitivity of initial learning, delayed recall, and the learning slopes ranged from 43.6-89.7 (Table 6).

## DISCUSSION

The presence of amyloid and APOEε4 allelic presence is each associated with AD, and early detection of these factors can open possibilities for intervention. Performance-based tests are a low cost and technologically portable method for potentially screening for these important biological prognostic factors. These results indicated that memory scores from the ANB have potential utility for detecting both APOEε4+ status and amyloid deposition. In stratifying by amyloid pathology, most total scores, including learning slope and delayed recall, approached adequate utility as a screening measure aside from initial learning did not distinguish between those with and without amyloid deposition. Most subtests also showed adequate utility, aside from delayed recall ALMT, RLS ALMT, and LR ALMT. A similar pattern of findings emerged when screening for APOEε4 status. In these analyses, each of the total scores and ANB subtest scores (aside from initial learning AVMT, RLS ALMT, and LOT ALMT), including initial learning, approached the threshold for being an adequate screen.

Research by Hammers and colleagues (2022) has suggested that the LR score has diagnostic advantages over other learning slope metrics such as RLS and LOT scores.^18^ In the present study, however, each learning slope score was roughly equivalent as a screening tool, with measures generally resulting in “fair” diagnostic properties and moderate differences in group means. The equivalence observed across measures is likely due to the lack of ceiling effects encountered in the present study compared to prior research that has demonstrated that ceiling effects are readily apparent in RLS scores when LR scores have been shown to be a superior measure over RLS scores. Our analyses revealed that the LR score was significantly correlated with initial learning, though shared a much stronger correlation with the other learning slope metrics that are dependent on raw improvement over the course of the test. The lack of ceiling effect essentially renders each of the learning slope scores as being interchangeable. In fact, no participant achieved a perfect score on the ALMT or AVMT.

Results also demonstrate that the ANB learning scores show promise in detecting the presence of amyloid and APOEε4status, but the diagnostic statistics fall just shy of justifying their use in isolation. Although these results do not provide strong support for using these measures independently, additional research is needed to examine whether they may provide useful information when combined with other variables. For instance, data from other tests or biomedical variables could be combined with ANB scores to provide a more refined screening for AD pathology. It is important to note that these results pertain to potentially subtle manifestations and/or risk factors for AD and were not applied to variables associated with more pronounced cognitive pathology, such as significant tau burden or neurodegeneration. We suspect that ceiling effects will be less problematic for advanced disease burden, and, therefore, each of the learning slope scores are likely to function interchangeably in this context as well. Clinically, additional research into understanding these learning metrics and cut-offs is warranted before using them in the SSA.

This study is the first in the SSA to attempt to predict the amyloid pathology and APOEε4status using cognitive screening measures (CSID and AQ), ANB memory subtests (ALMT and AVMT), and learning slope metrics (initial learning, RLS, LOT, and LR). This study has several limitations. First, this current study has a small sample of participants, which limited the detection of differences that could have been clinically and significantly relevant to discriminate the two groups. Thus, future studies should replicate these findings with larger sample sizes. Second, the screening measures used (CSID and AQ) have not been validated in the SSA/DRC. Third, this study included only subjects with suspected dementia and healthy controls. Those with cognitive difficulties seen in between the spectrum (e.g., MCI, subjective memory complaints) were excluded, leaving only the extremes of the dementia spectrum. Future studies should include all the 4 groups (healthy controls, MCI, subjective memory complaint and dementia). Additionally, this study analyzed only amyloid pathology and APOEε4 allele status. Other limitations include lack of normative data for the fluid biomarkers used here in the studied population, relatively small sample size for a biomarker study, and no replication cohort. Future studies should aim to replicate our findings along the AD pathology continuum, tau pathology, neurofilament light (NfL), as well as use other plasma biomarkers (e.g., glial fibrillar acidic protein [GFAP]), as they may provider greater insight into the progression of cerebral amyloid and tau pathology and cognitive decline in SSA populations. A major caveat is that amyloid positive was determined with plasma Aβ_42/40_ by Simoa, which is far from being the best biomarker for amyloid positivity. The gold standard is amyloid brain PET. Of the plasma biomarkers P-tau217 by mass spectrometry or ECL and Aβ_42_ / Aβ_40_ by mass spectrometry seem the best with AUC > 0.90. Plasma P-tau181 by Simoa is a reasonably robust alternative. It is not just a marker of tau pathology. It also is an accurate predictor of brain amyloid pathology detected by neuropathology of brain amyloid PET.^41^

## Data Availability

The data supporting the findings of this study are available on request from the corresponding author. The data are not publicly available due to privacy or ethical reasons.

